# ANTERIOR OR POSTERIOR ANKLE FOOT ORTHOSES FOR ANKLE SPASTICITY: WHICH ONE IS BETTER?

**DOI:** 10.1101/2021.03.02.21251574

**Authors:** Areerat Suputtitada, Watchara Chatkungwanson, Kittikorn Seehaboot, Carl PC Chen

## Abstract

**Backgroud:** Ankle foot orthoses (AFOs) have been widely used in stroke patients to assist in safe, energy efficient walking. Since they provide mediolateral ankle stability during stance, adequate toes clearance during swing phase and also facilitation of heel strike.

**Objective:** To compare the efficacy of anterior or posterior plastic ankle foot orthoses (AFOs) for ankle spasticity.

**Methods:** Crossover design with randomization for the interventions, blinded assessors was used. 20 patients with chronic stroke, Modified Ashworth scale (MAS) of ankle ≤ 2 and Tardieu angle ≥ 20 degrees. The participants were random to treat with anterior or posterior plastic AFOs and then were crossover to treat with the other one. Outcomes measurement was performed by using passive range of motion (PROM), MAS, walking velocity, stretch reflex surface EMG and walking surface EMG of medial gastrocnemius muscle. Patient satisfaction was evaluated at 1 month.

**Results:** Twenty stroke patients with ankle spasticity were recruited. Mean age was 46.60(38-60) years old.Mean duration. of times since stroke was 9.35 (6-15) months. Comparison between two types of orthoses revealed statistically significant improvement of walking surface EMG of medial gastrocnemius muscles when using anterior plastic AFO more than posterior plastic AFO at p=0.015. Patients satisfaction were statistically significant higher when using anterior plastic AFO more than posterior plastic AFO at p<0.05.

**Conclusion:** Anterior plastic AFO has more efficacy in reduced dynamic ankle spasticity during walking than posterior plastic AFO proven by comparison the dynamic electromyography changes in dynamic spasticity during walking.

## INTRODUCTION

Ankle foot orthoses (AFOs) have been widely used in stroke patients to assist in safe, energy efficient walking. They provide mediolateral ankle stability during stance, adequate toes clearance during swing and also facilitation of heel strike..^1-4^ Conventional plastic AFOs have a posterior leaf-type design, and are fabricated by a lamination or vacuum-forming technique over a positive plaster model of the limb.^1-7^ Anterior AFOs are low-temperature AFOs commonly used in Asian countries as they are light, easy to use, suitable for indoor walking and improve the postural stability.^8^ Prolonged stretching by a plastic ankle-foot orthosis is also used in spasticity management.^9^ Several studies have evaluated the effects of posterior AFOs on stroke patients and revealed improvement in gait parameters; including stride length, gait velocity and cadence, gait stability, balance control, energy cost of walking, and functional status.^1-7^ Several studies have evaluated the anterior AFO functions and suggested that anterior AFOs also work effectively for gait parameters, walking ability and balance control in hemiplegic stroke patients.^8,10,11^ Other studies compared the effect of anterior AFO with posterior AFO^10,11^ The superiority of one style over another is still inconclusive.^12^

Ankle motor control in stroke patients is variable and the designs of anterior AFOs and posterior AFOs are different. We speculated that anterior plastic AFO might have a superior efficacy in cases with predominately gastrocnemius and soleus spasticity since there would be less stimulation of spasticity due to direct contact of plastic materials. From the literature reviewed, this is the first study to compare the dynamic electromyography changes in static and dynamic spasticity of hemiplegic stroke patients using either anterior AFOs or posterior AFOs.

## MATERIALS AND METHODS

The Institutional Review Board of the Faculty of Medicine, Chulalongkorn University, Bangkok, Thailand. approved the study (IRB no. 47951). The clinical trial registry is TCTR20210225006.

Participants were informed about the procedure and provided written consent.

### Subjects

A randomized crossover clinical trial, blinded assessor was done to compare the effectiveness of anterior versus posterior plastic AFO for treatment of ankle spasticity in stroke patients. The inclusion criteria for the study group were as follows: (1) diagnosis of unilateral hemiplegia caused by either hemorrhagic or ischemic stroke; (2) ability to follow simple verbal commands or instructions; (3) Modified Ashworth scale (MAS) of ankle joints were ≤ 2 (4) Tardieu angles were ≥ 20 degrees (5) never have used an AFO and (6) ability to ambulate independently. Subjects were excluded if they had any of the following conditions: (1) medical problems other than stroke that would interfere with their gait; (2) foot-related premorbid or comorbid orthopedic problems; or (3) refused to be enrolled or sign the informed consent. All patients underwent neuroimaging studies, including computed tomography or magnetic resonance imaging of the brain, to confirm the diagnosis of stroke at an early stage.

### Instrumentation

The wireless surface EMG equipment (ME-6000, MEGA EMG) was used for quantitative measurement of spasticity.

### Procedure

A total of 124 stroke patients were assessed for eligibility. The Consolidated Standards of Reporting Trials (CONSORT) is shown in Figure 1., 104 of the patients were excluded. Of the remaining 20 patients were random to treat with anterior plastic AFO (Figure 2) or posterior plastic AFO (Figure 3). They applied each AFO for 30 days,6 hours per day. The AFO was removed for a 1-week interval before crossover to the other style. Every patient put on the same standard sandal shoes with Velcro strap. The AFOs were made of thermoplastic material and covered the mediolateral ankle joints all the time during walking. Outcomes measurement was performed by using passive range of motion (PROM), MAS, walking velocity, stretch reflex surface EMG and walking surface EMG of medial gastrocnemius muscle during terminal stance phase. Root Mean Square (RMS) of surface EMG were used as the indicators with Wireless surface EMG equipment (ME-6000, MEGA EMG®). **The** patient satisfaction was evaluated at the end of 30 days wearing before put off each AFO with a blinded assessor.

**Figure 1.**
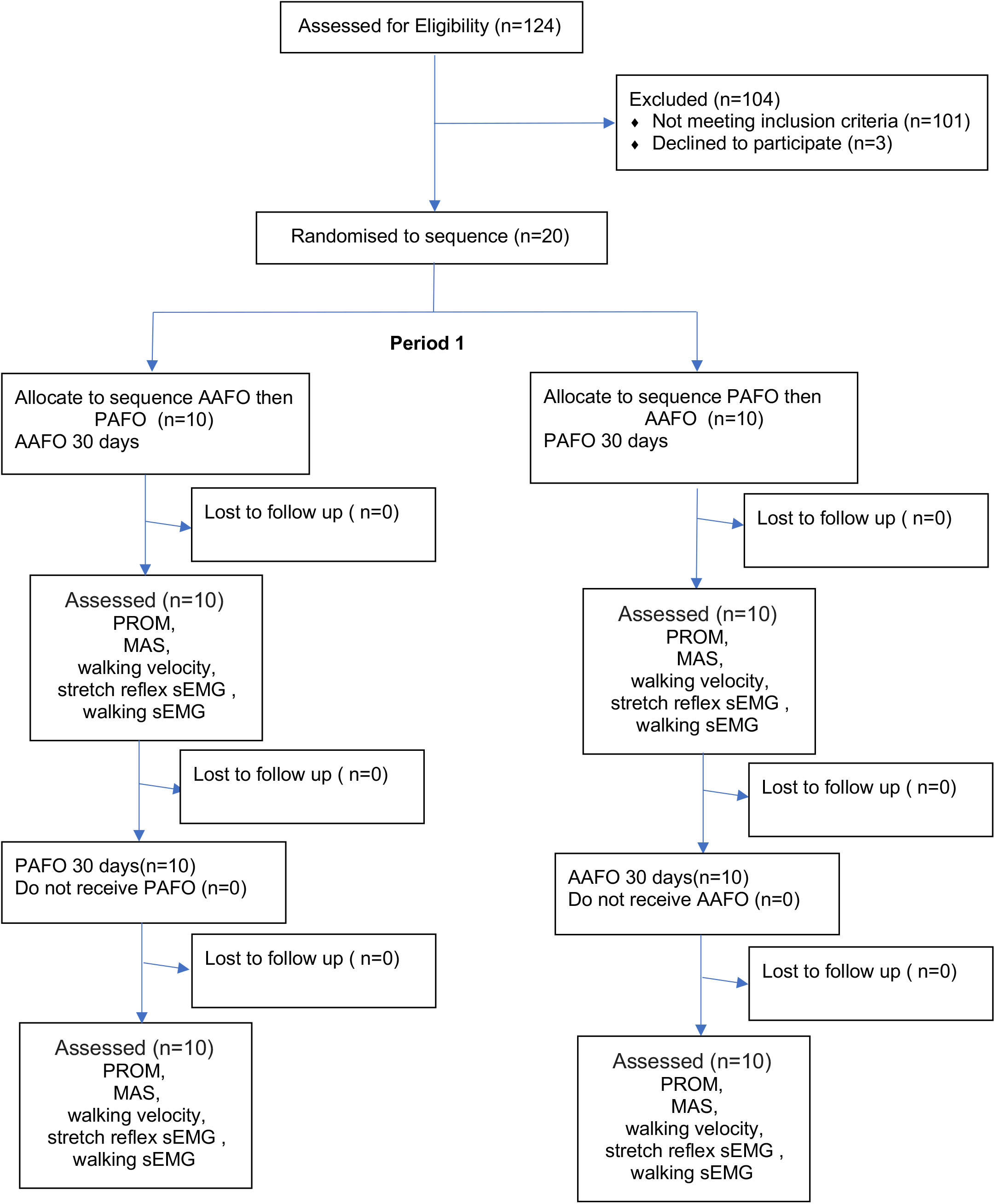
Consort Flow Chart.

**Figure 2.**
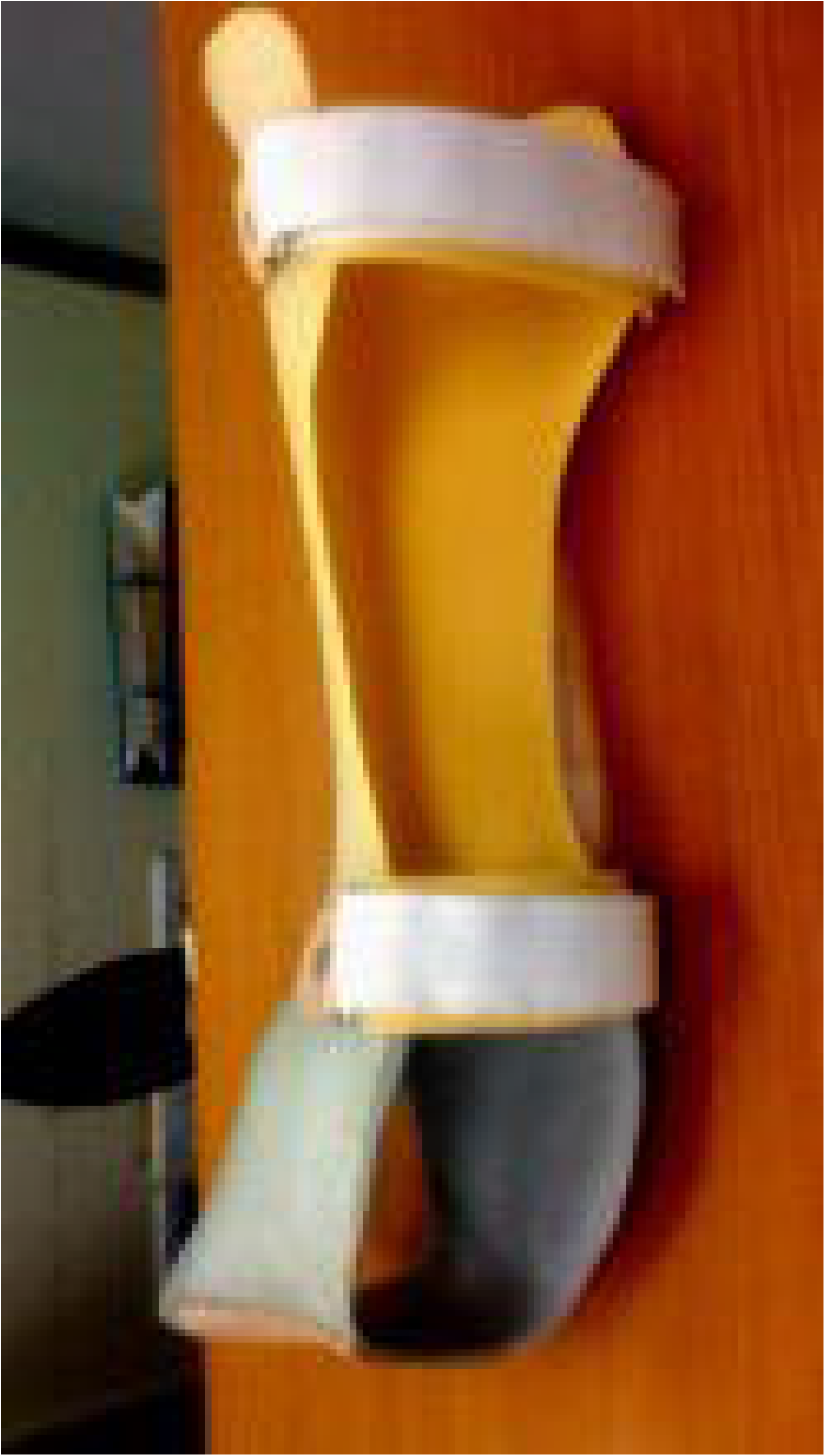
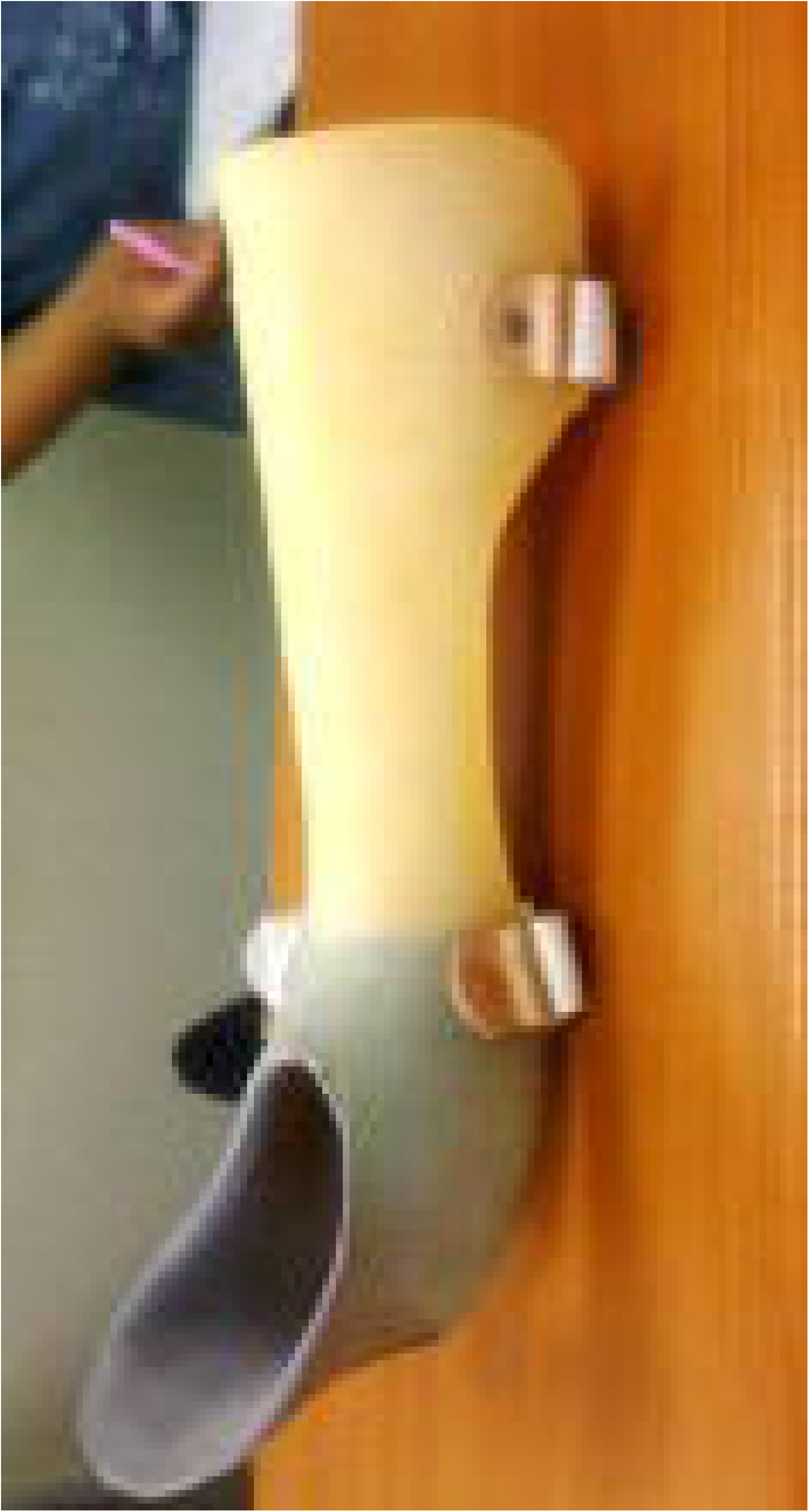
Anterior plastic AFO.

**Figure 3.**
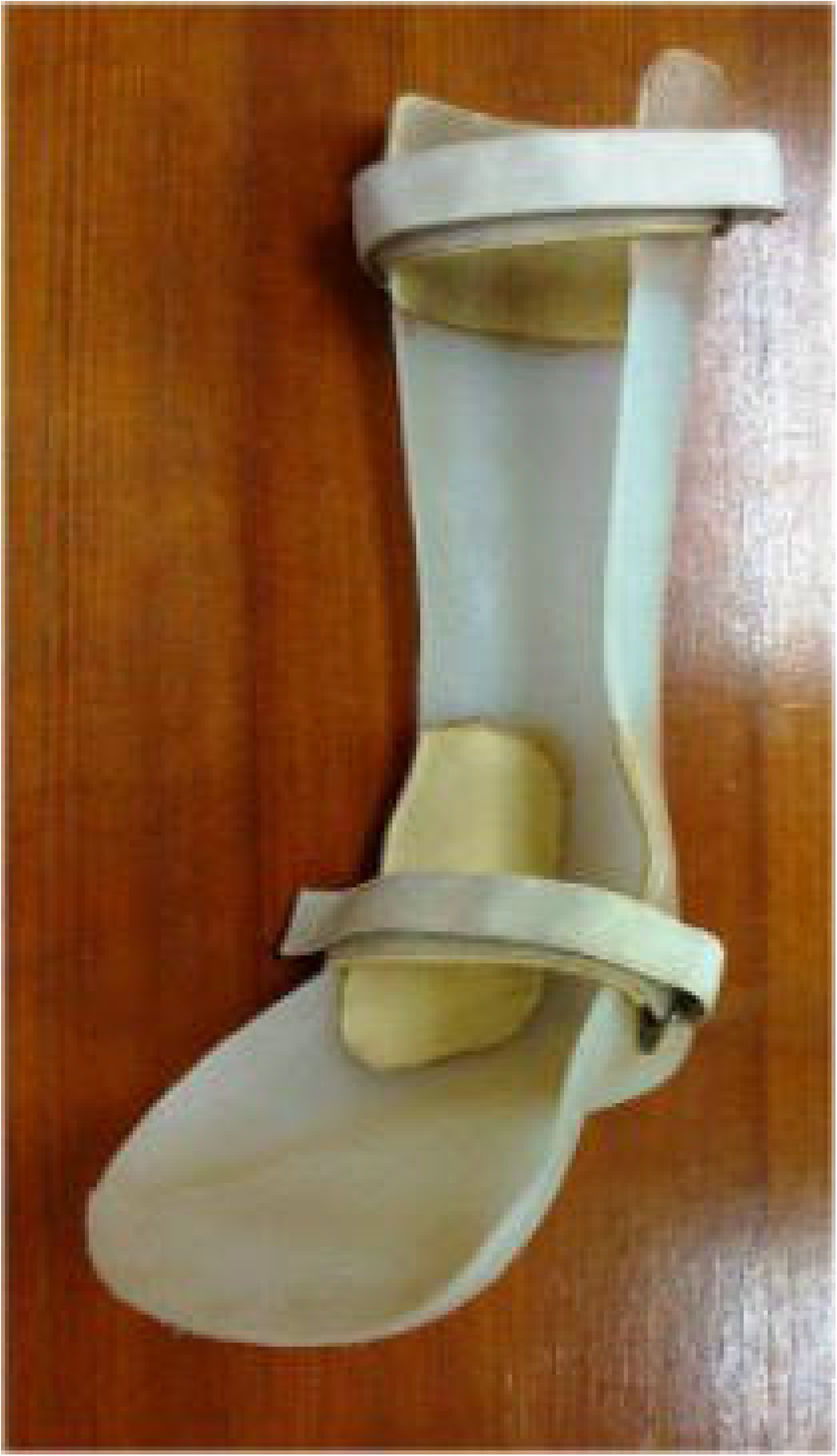
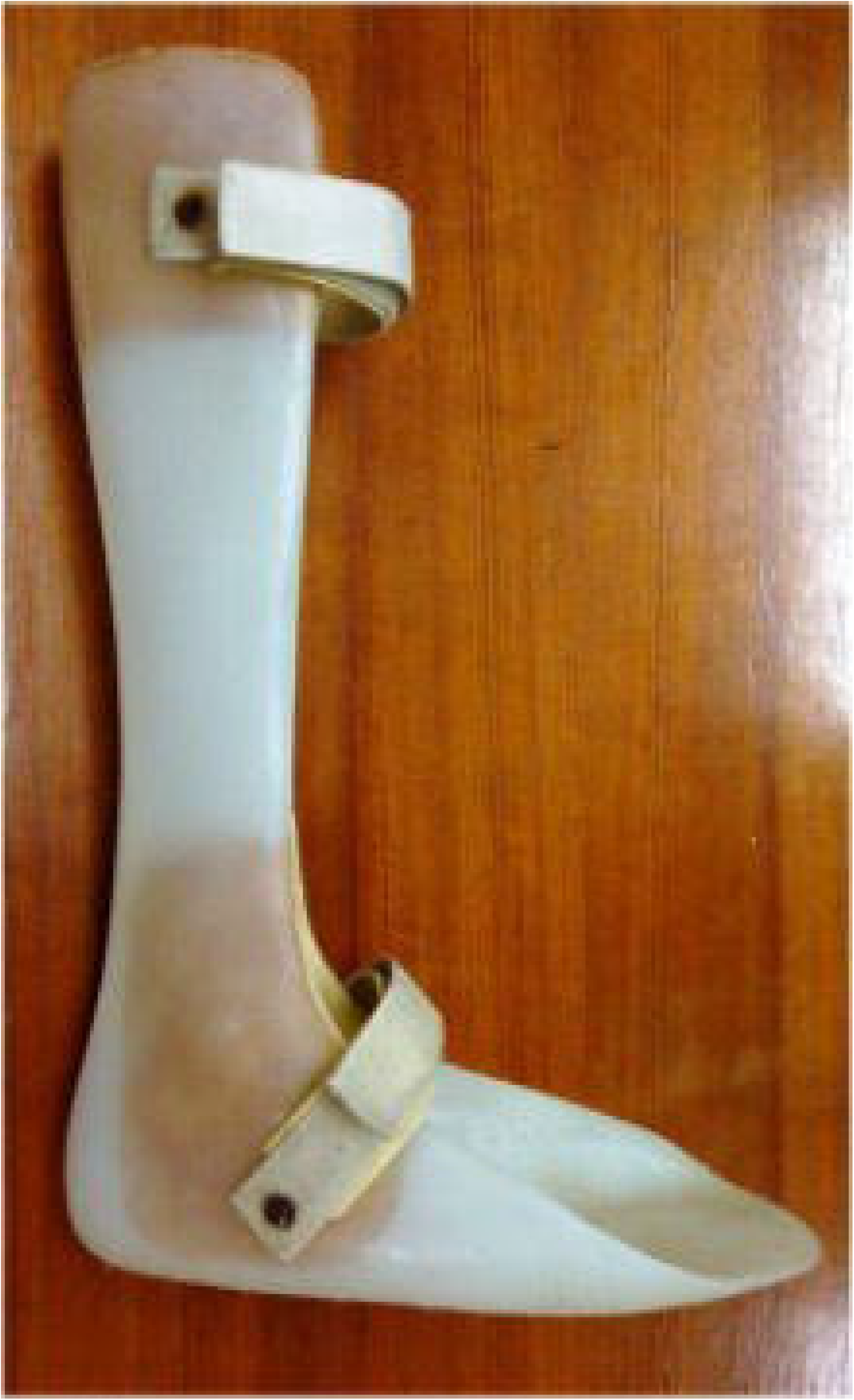
Posterior plastic AFO.

### Surface EMG measurement

The sEMG acquisition system consisted of a 6 Hz Butterworth low-pass flter to remove the noise of the sEMG signals, and the sEMG signals resolution a 14-bit analog to digital converter (ADC). The Stretch reflex RMS amplitude of Gastrocnemius (µV) ^13^ and Walking RMS amplitude of Gastrocnemius (µV) by sEMG were the mean of 3 times measurement.

### Data Analysis

All data were statistically analyzed using the Statistical Program for Social Sciences (SPSS) version 17 (SPSS Inc, Chicago). Descriptive statistics were displayed with mean, range, and standard deviation. Normality tests were done by Kolmogorov-Smirnov test. Intergroup comparisons were done with Mann Whitney U test, and the comparison within each group was tested using Wilcoxon signed-rank test. The level of significance used was *P* less than .05.

## RESULTS

Twenty stroke patients with ankle spasticity, MAS score at level ≤ 2 were recruited. Ten of them (50%) were male and Ten (50%) were female. Mean age was 46.60(38-60) years old. The mean time since onset of stroke was 9.35 (6-15) months. 12 of them (60%) were ischemic stroke from infarction and 8 of them (40%) were hemorrhagic stroke. Ten of them (50%) were randomized to use anterior AFO first and ten (50%) were random to use posterior AFO first and then were crossover to the other. Comparison between before and after using both types of AFO revealed statistically significant improvement of Passive range of motion (PROM), Modified Ashworth scale (MAS), walking velocity, stretch reflex and walking surface EMG of medial gastrocnemius muscles at terminal stance phase at p<0.05, as table 1. Comparison between two types of orthoses revealed statistically significant improvement of walking surface EMG of medial gastrocnemius muscles at terminal stance phase when using anterior plastic AFO more than posterior plastic AFO at p<0.05 as table 1. Patients satisfaction were statistically significant higher when using anterior plastic AFO more than posterior plastic AFO at p<0.05 as table 2.

**Table 1.**
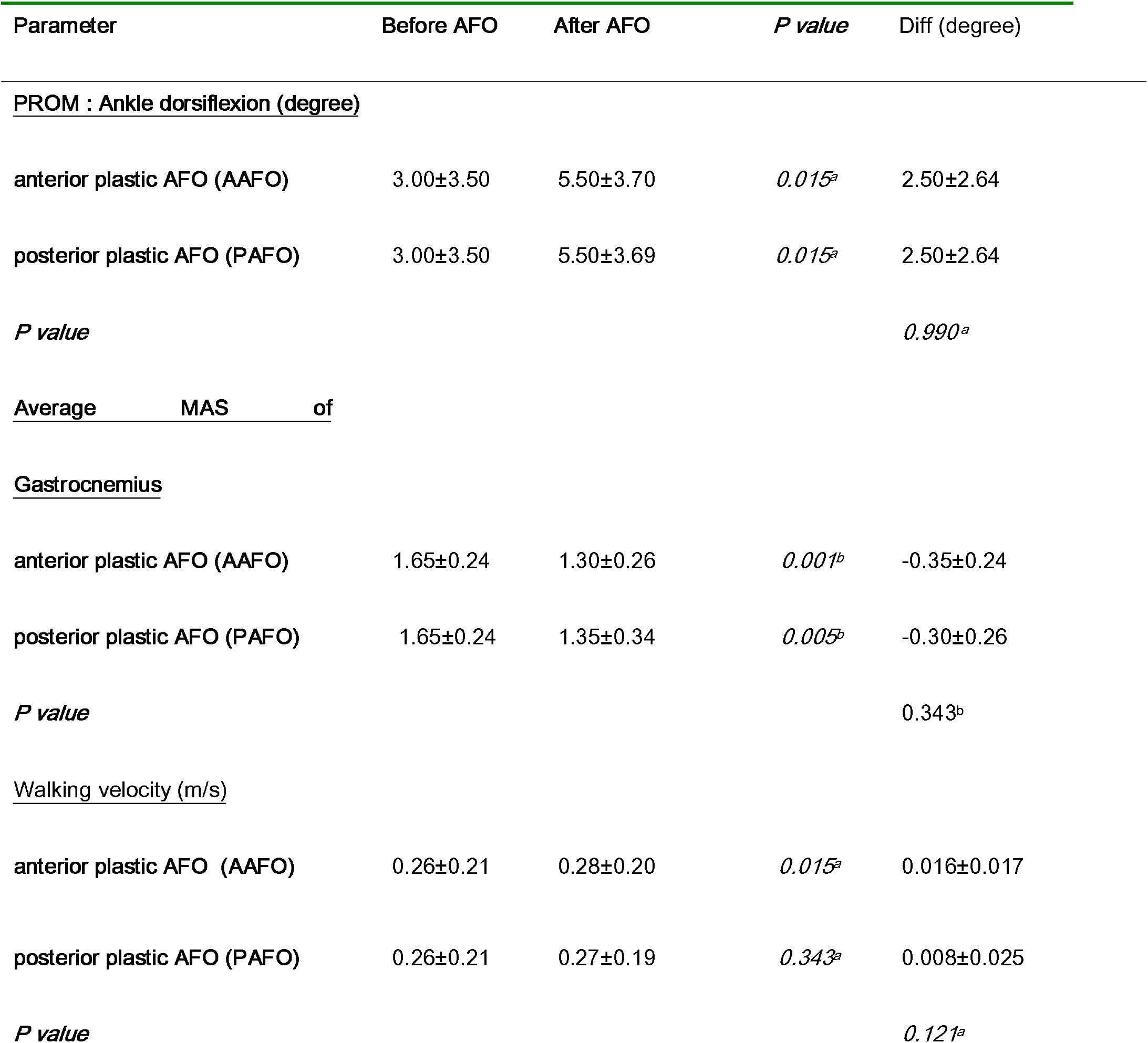
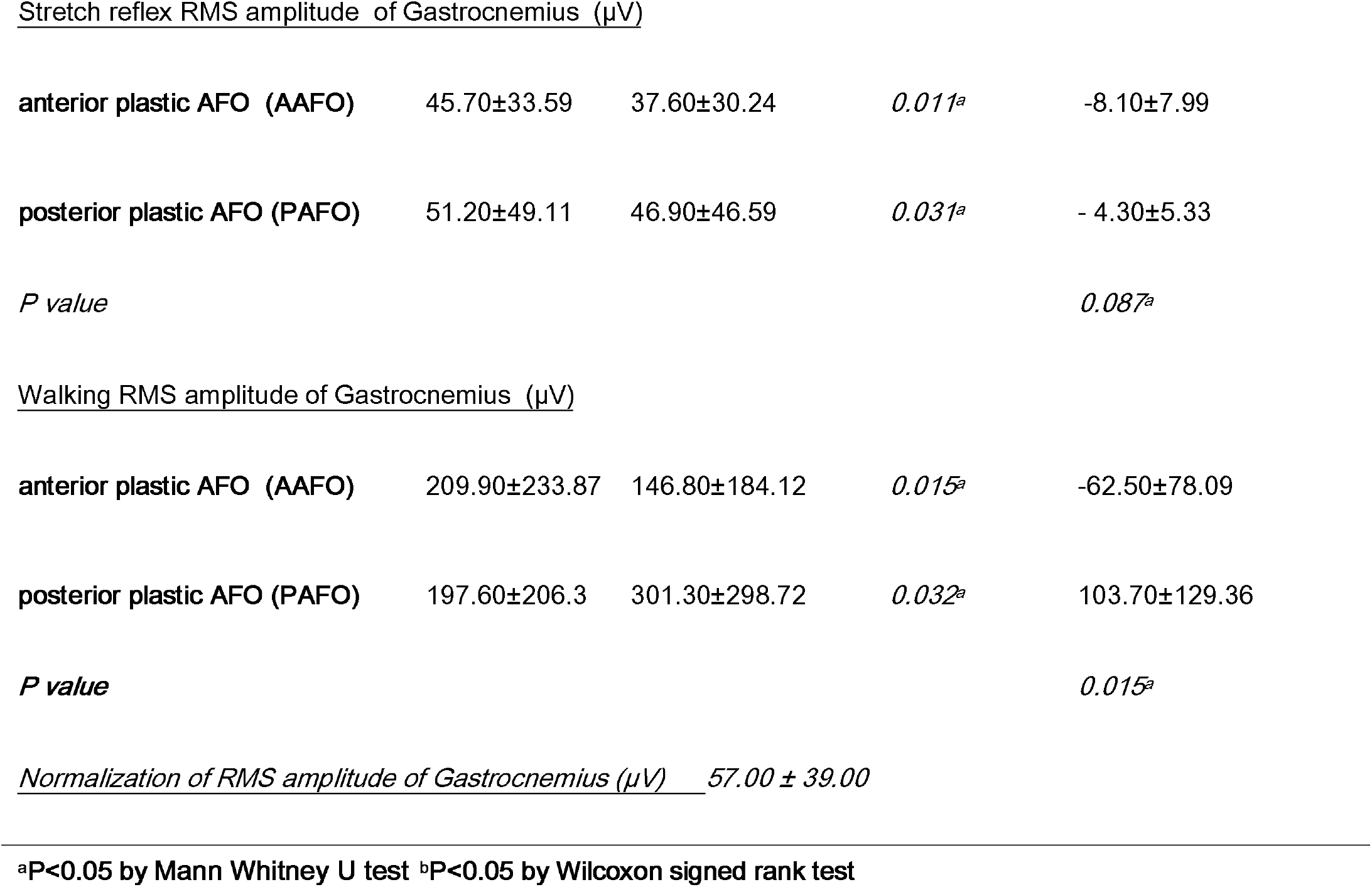
Compare between anterior plastic AFO (AAFO) and posterior plastic AFO (PAFO)

**Table 2.** Compare of patients’ satisfaction between anterior plastic AFO (AAFO) and posterior plastic AFO

| Parameter | AAFO | PAFO | *P value* |
| --- | --- | --- | --- |
| Beautiful | 3.09± 0.30 | 2.55± 0.52 | *0.006<sup>a</sup>* |
| Strength and durable | 2.91± 0.54 | 3.00± 0.00 | *0.341<sup>a</sup>* |
| Easy to put on | 2.09± 0.55 | 2.73± 0.47 | *0.046<sup>a</sup>* |
| Comfort during walking | 3.00 ± 0.00 | 2.54± 0.52 | *0.016<sup>a</sup>* |
| Reduce ankle spasticity | 3.00± 0.00 | 2.27± 0.47 | *<0.001<sup>a</sup>* |

<sup>a</sup>P<0.05 by Mann Whitney U test

## DISCUSSION

Dynamic Surface EMG measurement of stretch reflex amplitude was directed related to the MAS and RMS amplitude of muscle activity during walking, can be used to measure the dynamic spasticity.^13,14^ The result of surface EMG in this study showed anterior plastic AFO can reduce dynamic spasticity of gastrocnemius muscle during walking, and in addition increase walking speed more than posterior plastic AFO. The hypothesis was posterior plastic AFO may stimulate dynamic spasticity from contact surface of agonist muscles during walking. There is some evidences of spasticity inducement by stimulation of agonist muscles or ball of foot or palmar surface of hand. ^9^ In addition, the walking velocity when using the anterior plastic AFO was revealed to be significantly faster. This might be the effect of the ground reaction force with direct heel contact to the floor and the more ankle control to be dorsiflexion of anterior plastic AFO facilitating heel strike and rapid midstance with toe off.^15^ In this study all the patients have good heel strikes with their AAFO.

The Modified Ashworth Scale revealed a positive correlation with the amplitude and duration of Dynamic surface EMG response in stretch reflex maneuver as the static spasticity.^14^ The walking root mean square (RMS) amplitude and duration should be correlated with dynamic spasticity when walking. Our results revealed the statistically significant decrement of stretch reflex RMS amplitude and walking RMS amplitude of Gastrocnemius when using the anterior plastic AFO compared to baseline without using AFO and compared with using posterior plastic AFO. Whereas, there was no statistically significant decrement of stretch reflex RMS amplitude and moreover, increment of walking RMS amplitude when using the posterior plastic AFO. Our results supported the hypothesis of stimulation of spasticity by contact of plastic materials, especially when walking with more shearing force and stimulation of gastrocnemius spasticity by posterior plastic AFO. This hypothesis was tested and validated by Mayer et al regarding to the evidences of spasticity inducement by stimulates at agonist muscles or ball of foot or palmar surface of hand. ^9^ Chen CK et al revealed that in the early stage of recovery, the use of anterior plastic AFO may assist the improvement of postural stability. ^8^ The available evidences included all designs of AFO, cautiously suggested that an AFO can reduce energy cost, enhance weight transfer over the weak leg and improve ankle and knee kinematics in hemiplegic stroke patients.^15-18^ However, many clinicians and patients decline to prescribe, or use an AFO as many users complain about the weight, discomfort, difficulties fitting into shoes, or the appearance.^16-18^There are also fears that reliance on an AFO may induce muscle disuse and delay functional recovery.^16-18^ There were also insufficient common data to analyses the effect on ankle, knee and hip kinetics and muscle activity or spasticity. ^16-20^ Our result is the first research comparing the efficacy of anterior plastic AFO and posterior plastic AFO, which specify the measurement of spasticity relief by using outcome measure by novel of stretch reflex RMS amplitude and walking RMS amplitude of surface EMG at gastrocnemius muscles.

Failure to improve gait velocity in the group with posterior AFOs is contrary to the previous studies ^21-23^ The possible new finding should be addressed for any potential disadvantages of posterior AFO are as the followings; (1) the plastic full heel cord and plantar surface covering triggers muscle stretch reflex and increases muscle activities. ; (2) the solid AFO limit midfoot and forefoot movements by foot plate will inhibit the late stance phase if wearing shoe without rocker bottom. In contrast, it can be the advantage of anterior AFOs in patients who do not wear proper shoes with rocker mechanism or bare feet at home since their own feet can facilitate the late stance phase. Our patients had not used any orthoses previously, so that there is no potential bias (learning effect).

Onishi et al. ^23^ studied the relationship between dynamic EMG and muscle tension and found that during voluntary contraction, dynamic EMG and muscle tension show a certain level of positive correlation. Xue Ping L et als.^22^ also reported that dynamic surface EMG combined with isokinetic test is a reliable quantification means to assess the spasticity. Dynamic surface electromyography and isokinetic assessment can be used to estimate muscle spasticity of stroke patients in clinic. For the instrumented assessment, root mean square (RMS) electromyography and torque were better predictors for a positive response by logistic regression analysis (area under the ROC curve = .82), when compare to modified Tardieu angle. (area under ROC curve = 0.7)^22^ Hu B,et als used RMS of Stretch reflex by surface EMG to objective measurement of spasticity too.^13^

In our study, we also did a comparison of patient satisfaction between anterior plastic AFO and posterior plastic AFO and revealed statistically significant more satisfaction in comfort during walking and reduced spasticity but less satisfaction in the ease of donning when using both types of AFO since our anterior plastic AFO was designed to support mediolateral stability of ankle joints as well.

### Study limitation

The multidimensional nature of spasticity, reveals that there are different types of abnormal muscle activation during walking. The results of this study might be applicable to only the types of spasticity in our included patients. There are numerous studies revealed the efficacy of posterior plastic AFO.^1-7, 16-20.^ and anterior plastic AFO. ^8,10,11,15,17^ The explanation might need to specify many factors as the different designs of the AFOs, the time since onset of stroke, the types of spasticity as predominately gastrocnemius or tibialis posterior spasticity, any co-contracture of the soft tissues, etc. In the clinical practice, we may use either anterior or posterior plastic AFO and if the patients develop more spasticity, we can change to another type and anterior plastic AFO may be suitable during bare feet at home.

## CONCLUSION

Anterior plastic AFO has potential to reduce dynamic ankle spasticity during walking than a posterior plastic AFO. The anterior plastic AFO might be more effective in cases with predominately gastrocnemius and soleus spasticity since less stimulation of spasticity by contact of plastic materials proven by comparison the dynamic electromyography changes in dynamic spasticity during walking. The sEMG of stretch reflex and during walking were possible to be used for quantitative spasticity assessment. The further studies should be done to confirm.

## Data Availability

The data availability is at the Faculty of medicine,Chulalongkorn University

## Abbreviations

(AFO): Ankle-foot orthosis
(EMG): Electromyography
(PROM): Passive range of motion
(MAS): Modified Ashworth scale
(RMS): Root Mean Square

## Notes

### Competing Interest Statement

The authors have declared no competing interest.

### Clinical Trial

TCTR20210225006

### Funding Statement

Research grant from Medical Inventory and Technology Center, Chulalongkorn University.

### Author Declarations

The Institutional Review Board of the Faculty of Medicine, Chulalongkorn University,Bangkok,Thailand. approved the study (IRB no. 47951). Participants were informed about the procedure and provided written consent.

## REFERENCES

1. Wu KK. Ankle foot orthosis. In: Wu KK. Foot Orthoses: Principles and clinical applications. Baltimore: Williams&Wilkins. 1990:125–35.

2. Lehmann JF. Biomechanics of ankle foot orthosis: prescription and design. Arch Phys Med Rehabil. 1979;60:200–7.

3. Tyson SF, Thornton HA. The effect of a hinged ankle foot orthosis on hemiplegic gait:objective measures and users’ opinions. Clin Rehabil. 2001;15:53–8.

4. J.F. Lehmann, S.M. Condon, R. Price, B.J. deLateur Gait abnormalities in hemiplegia: their correction by ankle-foot orthoses. Arch Phys Med Rehabil. 1987; 68:763–71.

5. Leung J, Moseley A. Impact of ankle foot orthoses on gait and leg muscle activity in adults with hemiplegia:systematic literature review. Physiotherapy. 2003;89:39–55.

6. J.F. Lehmann, S.M. Condon, R. Price, B.J. deLateur Gait abnormalities in hemiplegia: their correction by ankle-foot orthoses. Arch Phys Med Rehabil. 1987; 68:763–71.

7. H. Abe, A. Michimata, K. Sugawara, N. Sugaya, S. Izumi. Improving gait stability in stroke hemiplegic patients with a plastic ankle-foot orthosis. Tohoku J Exp Med. 2009;218:193–9.

8. C.K. Chen, W.H. Hong, N.K. Chu, Y.C. Lau, H.L. Lew, S.F. Tang Effects of an anterior ankle-foot orthosis on postural stability in stroke patients with hemiplegia. Am J Phys Med Rehabil. 2008;87:815–20.

9. N.H. Mayer, R.M. Herman. Phenomenology of muscle overactivity in upper motor neuron syndrome. Eur Med Phys. 2004;40:85–110.

10. J.H. Park, M.H. Chun, J.S. Ahn, J.Y. Yu, S.H. Kang. Comparison of gait analysis between anterior and posterior ankle foot orthosis in hemiplegic patients. Am J Phys Med Rehabil. 2009;88:630–4.

11. A.M. Wong, F.T. Tang, S.H. Wu, C.M. Chen. Clinical trial of a low-temperature plastic anterior ankle foot orthosis. Am J Phys Med Rehabil. 1992;41–3.

12. Tyson SF1, Sadeghi-Demneh E, Nester CJ. A systematic review and meta-analysis of the effect of an ankle-foot orthosis on gait biomechanics after stroke. Clin Rehabil. 2013;27:879–91.

13. Hu B, Zhang X, Mu J, Wu M, Wang Y. Spasticity assessment based on the Hilbert-Huang transform marginal spectrum entropy and the root mean square of surface electromyography signals: a preliminary study. Biomed Eng Online. 2018 Feb 27;17(1):27. doi: 10.1186/s12938-018-0460-1.

14. Cooper A, Musa IM, van Deursen R, Wiles CM. Electromyography characterization of stretch responses in hemiparetic stroke patients and their relationship with the Modified Ashworth Scale. Clin Rehabil. 2005;19: 760 –6.

15. Chen CL, Teng YL, Lou SZ, Chang HY, Chen FF, Yeung KT Effects of an Anterior Ankle-Foot Orthosis on Walking Mobility in Stroke Patients: Get Up and Go and Stair Walking. Arch Phys Med Rehabil. 2014 Aug 12. pii: S0003-9993(14)00936-8. doi: 10.1016/j.apmr.2014.07.408.

16. Tyson SF, Kent RM. Effects of an ankle-foot orthosis on balance and walking after stroke: a systematic review and pooled meta-analysis. Arch Phys Med Rehabil. 2013;94:1377–85.

17. Chen C-C, Hong W-H, Wang C-M, et al. Kinematic features of rear-foot motion using anterior and posterior ankle-foot orthoses in stroke patients with hemiplegic gait. Arch Phys Med Rehabil 2010; 91: 1862–8.

18. Leung J, Moseley A. Impact of ankle-foot orthoses on gait and leg muscle activity in adults with hemiplegia: systematic literature review. Physiotherapy. 2003;89: 39–55.

19. Pohl M, Mehrholz J. Immediate effects of an individually designed functional ankle-foot orthosis on stance and gait in hemiparetic patients. Clin Rehabil. 2006; 20:324–30..

20. Franceschini M, Massucci M, Ferrari L, Agosti M, Paroli C. Effects of an ankle-foot orthosis on spatiotemporal parameters and energy cost of hemiparetic gait. Clin Rehabil. 2003; 17: 368–72.

21. Carse B, Bowers R, Meadows BC, Rowe P. The immediate effects of fitting and tuning solid ankle-foot orthoses in early stroke rehabilitation. Prosthet Orthot Int. 2015 Dec;39(6):454–62.

22. Xue Ping L., Kai C, Jun Z, et al. Clinical assessment of muscle spasticity in stroke patients with surface electromyography and isokinetic assessment. Chin J Mod Med. 2010;605-8.

23 Onishi H, Yagi R, Akasab K et al. Relationship between EMG signals and force in human vastus lateralis muscle using multiple bipolar wire electrodes. J Electromyogr Kinesiol. 2000; 10:59–67

